# Increased transmissibility of the B.1.1.7 SARS-CoV-2 variant: Evidence from contact tracing data in Oslo, January to February 2021

**DOI:** 10.1101/2021.03.29.21254122

**Authors:** Jonas Christoffer Lindstrøm, Solveig Engebretsen, Anja Bråthen Kristoffersen, Gunnar Øyvind Isaksson Rø, Alfonso Diz-Lois Palomares, Kenth Engø-Monsen, Elisabeth Henie Madslien, Frode Forland, Karin Maria Nygård, Frode Hagen, Gunnar Gantzel, Ottar Wiklund, Arnoldo Frigessi, Birgitte Freiesleben de Blasio

**Affiliations:** Division of Infection Control and Environmental Health, Norwegian Institute of Public Health, Oslo, Norway; SAMBA, Norwegian Computing Center, Oslo, Norway; Telenor Research, Oslo, Norway; Oslo Municipality Health Service, Oslo, Norway; Oslo Centre for Biostatistics and Epidemiology, Institute of Basic Medical Sciences, University of Oslo, Norway; Oslo Centre for Biostatistics and Epidemiology, Oslo University Hospital, Oslo, Norway

**Keywords:** SARS-CoV-2, transmissibility, B.1.1.7, household transmission, contact tracing data, secondary attack rate

## Abstract

We use data from contact tracing in Oslo, Norway, to estimate the new SARS-CoV-2 B.1.1.7 lineage’s relative transmissibility. Within households, we find an increase in the secondary attack rate by 60% (20% 114%) compared to other variants. In general, we find a significant increase in the estimated reproduction number of 24% (95% CI 0% - 52%), or an absolute increase of 0.19 compared to other variants.

---

New, more transmissible variants of the SARS-CoV-2 pose a significant challenge to the response against the ongoing Covid-19 pandemic. The B.1.1.7 lineage that emerged in the United Kingdom (UK) in September 2020 [1] and subsequently spread to other countries and is reported to be more transmissible than the previously circulating lineages [2, 3]. There is also evidence of higher risk of hospitalisation for the B.1.1.7 variant [4, 5]. Though our study is not the first to estimate the relative transmissibility of B.1.1.7, local estimates are imperative due to differences in infection control measures, population behaviour, and other circulating variants. Contact tracing data contain important information that can be used to estimate secondary attack rates (SAR) and reproduction numbers during an emerging epidemic [6].

Using data from the contact tracing database PasInfo in Oslo Municipality, Norway, we compared the number of secondary cases among close contacts of persons infected with B.1.1.7 with those infected with other circulating lineages to estimate the increased transmissibility of B.1.1.7 in Oslo.

The data contain information on primary cases that tested positive in early 2021, from January 4 until February 28. Primary cases were eligible for inclusion if the virus lineage test result was based on Whole-Genome Sequencing or Sanger Sequencing, and they were recorded as infected outside their household. For each primary case, the record contains age, number of close contacts, and number of SARS-CoV-2 positive close contacts within and outside the household. Close contacts are defined as persons who had direct physical or close contact (≥ 15 minutes and < 2 meter) within 48 hours of symptom onset of the case or time of test for asymptomatic cases.

During the study period, testing on arrival at the Norwegian border was obligatory, and all direct flights from the UK were cancelled for several weeks. In Oslo, social activities in homes were discouraged, cultural events were banned, teaching at universities was digital, teleworking was encouraged, restaurants could not serve alcohol, schools were in part digital except for children below 13 years. When the first non-imported cases of B.1.1.7 were detected on January 23, all non-essential shops closed, and teleworking was obligatory when possible. Some of these restrictions were eased on February 16 [7].

## Results

After exclusion of 45 cases with incomplete information, the contact tracing data included 415 index cases, and their 2718 reported close contacts, of which 368 tested positive. In total, 5818 positive cases were registered in Oslo in the same period. Table 1 show the demographic composition of the sequenced primary cases. 146 of these were infected with the B.1.1.7 lineage, while 269 were infected with other lineages. For 251 of the cases (85 B.1.1.7, 166 other lineages) at least one close contact within her/his household was recorded and included in the analysis of household transmission.

**Table 1:** The number of primary cases and their average number of close contacts and secondary infections by age and lineage

|  | B.1.1.7 | Other lineages |
| --- | --- | --- |
| Number of primary cases per age group |  |  |
| 0-19 years | 14 | 28 |
| 20-39 years | 68 | 124 |
| 40-59 years | 47 | 73 |
| 60+ years | 16 | 44 |
| Total | 146 | 269 |
| Mean number of close contacts within households (sd) |  |  |
| 0-19 years | 2.79 (1.63) | 2.70 (1.56) |
| 20-39 years | 1.03 (1.36) | 1.38 (1.60) |
| 40-59 years | 1.72 (1.87) | 1.45 (1.43) |
| 60+ years | 1.25 (1.65) | 0.34 (0.64) |
| All ages | 1.45 (1.67) | 1.36 (1.55) |
| Mean number of secondary infections within households (sd) |  |  |
| 0-19 years | 0.86 (0.95) | 0.63 (1.15) |
| 20-39 years | 0.31 (0.63) | 0.31 (0.73) |
| 40-59 years | 1.09 (1.48) | 0.48 (0.75) |
| 60+ years | 0.38 (0.81) | 0.11 (0.32) |
| All ages | 0.61 (1.07) | 0.36 (0.75) |
| Mean number of close contacts (sd) |  |  |
| 0-19 years | 19.64 (26.01) | 15.00 (22.33) |
| 20-39 years | 5.30 (7.22) | 7.01 (18.86) |
| 40-59 years | 6.09 (6.72) | 4.71 (5.09) |
| 60+ years | 4.38 (4.86) | 2.00 (2.56) |
| All ages | 6.83 (10.94) | 6.40 (15.28) |
| Mean number of secondary infections (sd) |  |  |
| 0-19 years | 1.07 (1.27) | 1.36 (1.54) |
| 20-39 years | 0.61 (0.77) | 0.73 (1.33) |
| 40-59 years | 1.51 (1.63) | 0.90 (1.42) |
| 60+ years | 1.25 (1.61) | 0.59 (0.95) |
| All ages | 1.01 (1.30) | 0.82 (1.34) |

Among those infected with the B.1.1.7 lineage, the average number of close contacts was 6.83 (standard deviation (SD): 10.9), and the average number of secondary infections was 1.01 (SD: 1.3). Within the household, the average number of close contacts was 1.45 (SD: 1.7), and the average number of secondary infections was 0.61 (SD: 1.1).

Among those infected with the other variants, the average number of close contacts was 6.4 (SD: 15.3), and the average number of secondary infections was 0.82 (SD: 1.3). Within the household, the average number of close contacts was 1.36 (SD: 1.6), and the average number of secondary infections was 0.36 (SD: 0.8). The total number of close contacts of index cases in the age group < 19 years is higher likely due to cohorting and school class testing. The number of contacts is overdispersed, in accordance with previous findings from Ireland [8].

We used Poisson regression, with the number of secondary cases as the response to estimate the SAR and the relative risk and for obtaining age-adjusted estimates of these. To obtain the lineage-specific SAR, we fitted separate intercept-only models without covariates to the B.1.1.7 data and the other lineages data. The relative risk was estimated using an indicator variable for the cases infected with the B.1.1.7 lineage. We included an offset to account for the number of close contacts in all models; hence the SAR is the probability of secondary infection per close contact. Age group was added to the models using sum-to-zero constraints to preserve the interpretation of the intercept and of the relative risk parameters. Table 2 shows the estimated SAR for primary cases with the B.1.1.7 and other lineages, in addition to the ratio between them.

**Table 2:**
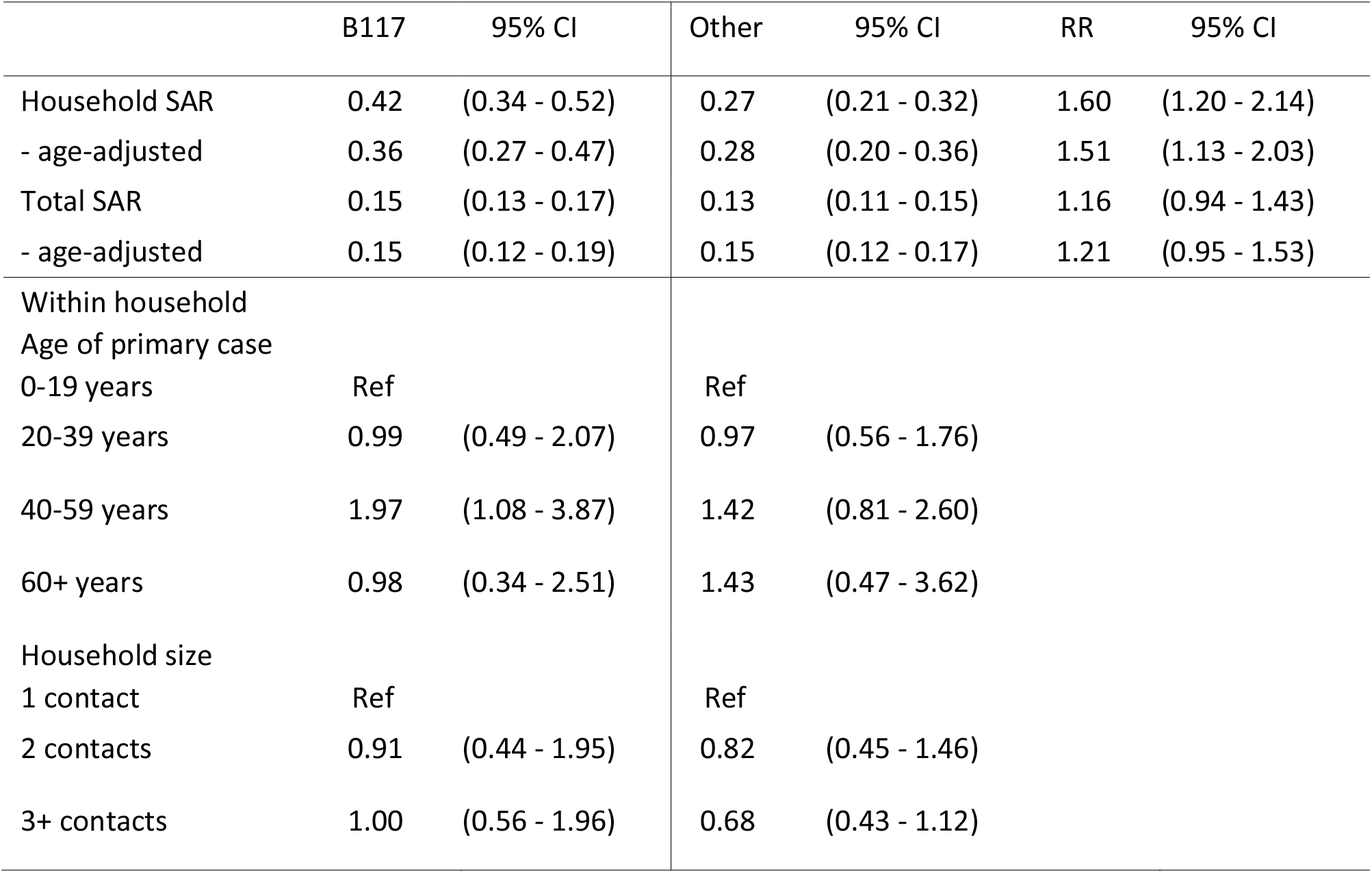
The estimated secondary attack rates (SAR) for the B.1.1.7 lineage and for other lineages, with the relative risk between them, and univariate analyses of the impact of age of primary case and household size on SAR within households for the B.1.1.7 variant and other lineages.

|  | B117 | 95% CI | Other | 95% CI | RR | 95% CI |
| --- | --- | --- | --- | --- | --- | --- |
| Household SAR | 0.42 | (0.34 - 0.52) | 0.27 | (0.21 - 0.32) | 1.60 | (1.20 - 2.14) |
| - age-adjusted | 0.36 | (0.27 - 0.47) | 0.28 | (0.20 - 0.36) | 1.51 | (1.13 - 2.03) |
| Total SAR | 0.15 | (0.13 - 0.17) | 0.13 | (0.11 - 0.15) | 1.16 | (0.94 - 1.43) |
| - age-adjusted | 0.15 | (0.12 - 0.19) | 0.15 | (0.12 - 0.17) | 1.21 | (0.95 - 1.53) |
| Within household |  |  |  |  |  |  |
| Age of primary case |  |  |  |  |  |  |
| 0-19 years | Ref |  | Ref |  |  |  |
| 20-39 years | 0.99 | (0.49 - 2.07) | 0.97 | (0.56 - 1.76) |  |  |
| 40-59 years | 1.97 | (1.08 - 3.87) | 1.42 | (0.81 - 2.60) |  |  |
| 60+ years | 0.98 | (0.34 - 2.51) | 1.43 | (0.47 - 3.62) |  |  |
| Household size |  |  |  |  |  |  |
| 1 contact | Ref |  | Ref |  |  |  |
| 2 contacts | 0.91 | (0.44 - 1.95) | 0.82 | (0.45 - 1.46) |  |  |
| 3+ contacts | 1.00 | (0.56 - 1.96) | 0.68 | (0.43 - 1.12) |  |  |

Within households, the estimated SAR is 0.27 for the non-B.1.1.7 lineages, and 0.42 for the B.1.1.7 lineage. The B.1.1.7 lineage is estimated to be 60% (20% - 114%) more transmissible within households than the other circulating variants. Index cases aged 40-59 years had a significantly higher risk of infecting household members if infected with the B.1.1.7 lineage. Household size was not found associated with higher SAR. Independently of the location, the estimated SAR is 0.13 for the non-B.1.1.7 lineages, while it is 0.15 for the B.1.1.7 lineage. The B.1.1.7 lineage is estimated to be 16% (−6% - 43%) more infectious than the other circulating variants. Adjusting for the age of the primary cases did not substantially change the estimates.

We used the same Poisson regression approach to estimate reproduction numbers but without adjusting for the total number of contacts. The results are shown in Table 3. The reproduction number for the B.1.1.7 lineage is estimated to be 24% greater than for the other lineages, or 0.19 difference in absolute numbers.

**Table 3:** Estimated reproduction numbers for the B.1.1.7 lineage and for other lineages, as well as the relative ratio between them.

| Reproduction number | Estimate | 95%CI |
| --- | --- | --- |
| R other lineages | 0.82 | (0.72 – 0.93) |
| R B.1.1.7 | 1.01 | (0.86 - 1.19) |
| Ratio | 1.24 | (1.00 - 1.52) |

## Discussion and conclusion

We estimate a 60% higher transmissibility of lineage B.1.1.7 within households. To our knowledge, this is the first estimate of the relative transmissibility of the B.1.1.7-variant in household settings. Our estimate is compatible with results from the United Kingdom that report higher transmissibility in the 43%–82% range [2] and 40%-80% [9]. Our findings are also in accordance with results from Denmark, which report 36% higher transmissibility for the B.1.1.7 variant than the other circulating Danish variants [3]. Note that our 95% confidence interval is wide (20% - 114%).

When we also consider contacts outside households, we find lower relative transmissibility. There are two potential explanations for this. There could be bias in the data when considering all close contacts. By restricting our analysis to contacts within the household, we control for potential selection bias of contacts introduced by the contact tracing, as contacts within households are more likely to be complete in any case and thus comparable across variant. Another possibility is that there is very little difference in the transmissibility for contacts outside households because of stringent and efficient distancing interventions. Transmission in the home setting has been shown to be more important during strong interventions [10]. The fact that the estimates are not affected when we control for age means that the susceptibility by age group is similar for B.1.1.7 and for the other circulating lineages. This finding is in line with studies from other countries, e.g. from UK showing that B.1.1.7 is associated with increased transmissibility in all age groups [11].

We estimated a household attack rate of 27% for non-B.1.1.7 lineages, which is lower than 45%, reported in a Norwegian prospective household study enrolling cases between February and April 2020 [12]. As testing was not easily available during the early phase of the pandemic, delay in testing may have increased the exposure time for household members, thereby contributing to the high transmission observed in this study. Still, both estimates are high compared to 17%, found in a systematic review of studies of the spread of SARSCoV-2 within households; however, documenting high heterogeneity between studies [13].

The data are subject to several limitations. We cannot distinguish between the case infecting the contact or vice versa. One contact could be the contact of several cases. There can be undetected cases: close contacts who are not registered, infected contacts that escape detection, and infected contacts who have not yet developed the infection (censored). Some non-positive contacts could be immune due to a previous infection. However, we do not expect any of these problems to occur often for some lineages than others, and hence the relative estimates should be less affected by these sources of bias.

Besides, there are potential sources of bias in the contact tracing data, which could affect our estimates. Even though the definition of close contact and contact tracing instructions were the same for the different lineages, there were likely differences in how the tracing was performed in practice, with an increased focus on tracing and testing contacts of cases infected with the B.1.1.7 variant. Due to the increasing circulation of B.1.1.7 in Oslo, the TICQ (Test-Isolation-Contact tracing-Quarantine) intervention was strengthened. In addition to the general requirement of quarantine for all close contacts of confirmed SARSCov-2 cases, since 5 February, all close contacts were tested at the beginning and end of their quarantine and household members of close contacts of cases confirmed with lineage B.1.1.7 were asked to quarantine until the contact was confirmed negative. Since March, quarantining the household members of close contacts was extended to include all close contacts regardless of the variant. The expanded TICQ targeting lineage B.1.1.7 cases may have introduced bias on the number of contacts registered outside the home, depending on lineage, thereby affecting our relative SAR estimate based on the total number of contacts. However, it should not affect the relative household SAR, as household members are known and followed up regardless.

Our study suggests that households are major locations for rapid transmission of the lineage B.1.1.7. Therefore, lowering the risk of spread within the families is pivotal to controlling the covid-19 pandemic with new and more transmissible lineages, given the Norwegian infection control policy with expansive household quarantining. However, it is strenuous to avoid transmission in households and more difficult under crowded conditions, potentially introducing a socioeconomic gradient [14]. The Norwegian government is now proposing a more extensive use of quarantine hotels related to inbound travel. This may also be encouraged for domestic cases with more transmissible virus variants, particularly in situations of unvaccinated household members in the risk groups.

## Data Availability

The data is person sensitive and cannot be shared publically.

---

**Figure 1:**
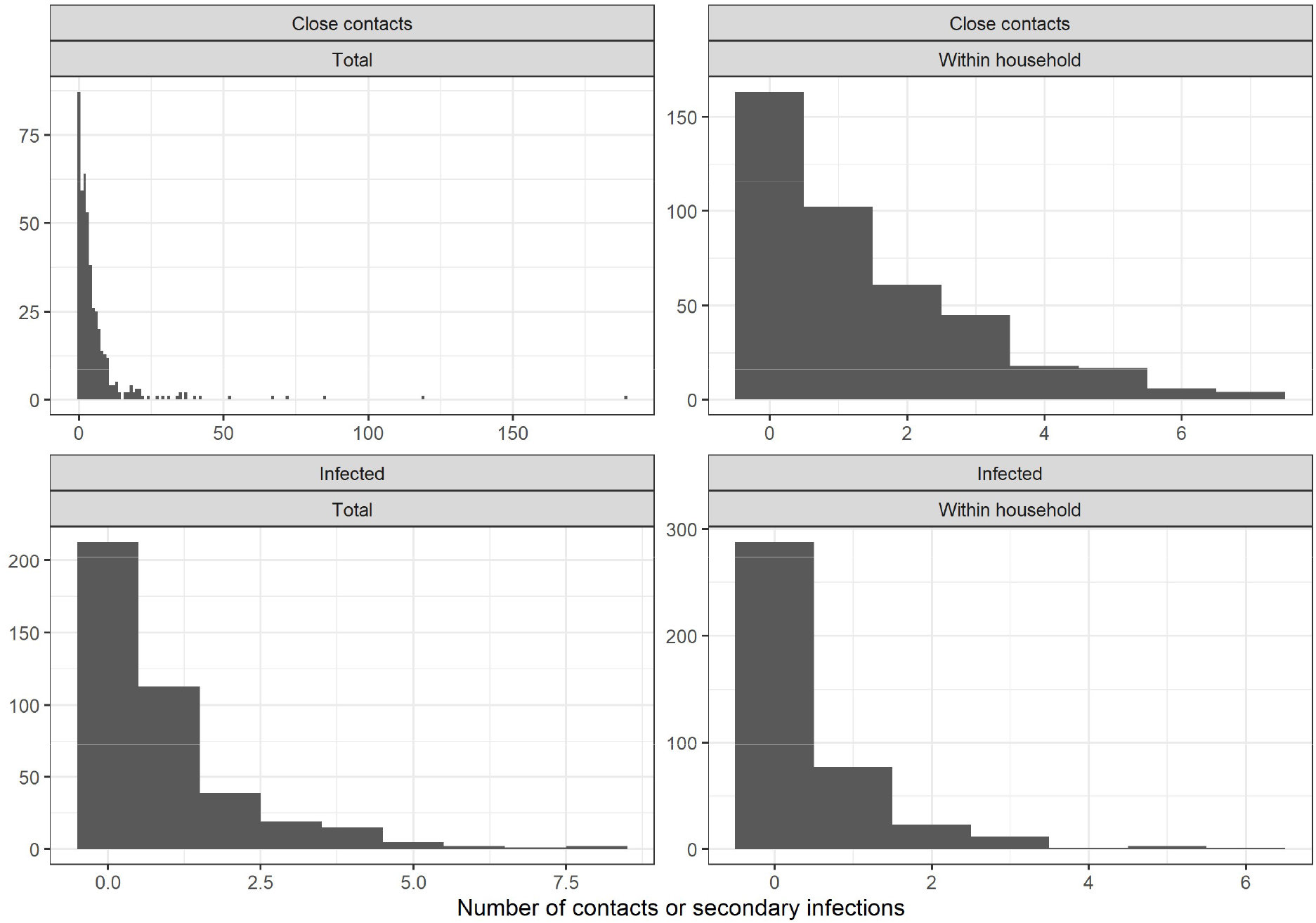
The distribution of the number of close contacts (top) and the number of secondary cases (bottom), in total (left), and within households (right) of all lineages.

